# Causal Effects of Herpesvirus-Associated Antibodies on Autoimmune neuroinflammatory diseases: Mendelian Randomization Study

**DOI:** 10.1101/2024.10.31.24316542

**Authors:** Shujun Sun, Yiyong Wen, Tang Pan, Xie Fan, Wen Jun, Anding Xu

## Abstract

**Background:** Herpesvirus infections may trigger Autoimmune neuroinflammatory diseases (ANDs), but their causal role is uncertain. This study used Mendelian randomization (MR) to investigate the causal effects of herpesvirus antibodies on ANDs.

**Methods:** We assessed 22 herpesvirus antibodies and five ANDs—multiple sclerosis (MS), neuromyelitis optica (NMO), myasthenia gravis (MG), Guillain-Barr é syndrome (GBS), and chronic inflammatory demyelinating polyneuropathy (CIDP)—using five MR methods. MR-PRESSO, MR-Egger, and Cochran’s Q statistic identified pleiotropy and heterogeneity. Bonferroni correction set the significance threshold at P<4.545e-04. Validation used UK Biobank and FinnGen data, with meta-analysis for inconsistent results.

**Results:** Significant positive causal effects were found for Epstein-Barr virus encoded nuclear antigen-1 (EBV EBNA-1) (OR=3.009), EBV viral capsid antigen (VCA) p18 (OR=8.700), and human herpesvirus 7(HHV7) U14 (OR=3.359) antibody levels on MS risk, and HHV7 U14 (OR=10.641), Varicella zoster virus glycoprotein E and glycoprotein I(VZV gE and gI) (OR=17.220) antibody levels on NMO risk. VZV gE and gI (OR=2.542) antibodies increased MG risk, while EBV ZEBRA (OR=0.592)reduced MS risk. Negative effects of EBV EA-D and ZEBRA antibodies on NMO were also noted. Validation confirmed the effects of EBV EBNA-1, EBV VCA p18 antibodies on MS, and VZV gE and gI on antibody MG, while EBV ZEBRA and HHV7 U14 antibodies effects on MS were not validated.

**Conclusions:** The study suggests a causal link between specific herpesvirus antibodies and the risk of MS, NMO, and MG.

## Introduction

Autoimmune neuroinflammatory diseases (ANDs) are a category of disorders in which the immune system mistakenly targets and attacks the nervous system, resulting in inevitable structural and functional damage[1, 2]. Autoimmune neurological diseases (ANDs) include mainly multiple sclerosis (MS), neuromyelitis optica (NMO), Guillain-Barr é syndrome (GBS), chronic inflammatory demyelinating polyneuropathy (CIDP) and myasthenia gravis (MG). MS and NMO affect the central nervous system (CNS), GBS and CIDP affect the peripheral nervous system (PNS), and MG affects the neuromuscular junction (NMJ). These conditions lead to neuronal or axonal damage, demyelination, damage to the nerve-muscle junction and other pathological changes [3, 4].

The incidence and prevalence of autoimmune diseases are increasing worldwide every year, but the exact causes of these diseases remain unknown; however, it is thought that a mixture of genetic and environmental factors play a role in their development [5]. Infections seem to be a common trigger for autoimmune diseases [6].

Several viruses of the Herpesviridae family have been implicated as an important risk factor for ANDs, Epstein-Barr virus (EBV) has been implicated as a major cause of multiple sclerosis [7], the risk of MS increases 32-fold following infection with EBV [8]. Human herpesvirus 6A (HHV-6A) has been associated with a higher risk of developing multiple sclerosis. Cytomegalovirus (CMV) infection may be associated with the occurrence of aquaporin-4 antibody-positive NMO spectrum disorder (AQP4 IgG+ NMOSD) [9]. CMV and EBV are also frequently identified as antecedent pathogens of GBS [10]. Case reports of CIDP have found associations with CMV and EBV [11].

The association between EBV infection and the pathogenesis of MG remains controversial [12, 13]. Meanwhile, the associations between viruses of the Herpesviridae family and MG remain unclear. Existing observational studies and case reports have limited sample sizes, which reduces the statistical power to detect true associations and increases the likelihood of random error. In addition, it is difficult to draw causal conclusions from observational evidence due to residual confounding and reverse causation [14].

Mendelian randomisation (MR) is a novel approach that may offer more effective solutions to the above shortcomings. MR uses genetic variations that affect exposure as instrumental variables (IVs) to assess the causal relationship between exposure and outcomes. Because alleles are randomly assigned at conception, they are unaffected by confounding factors and disease states. By using genetic variants as IVs, MR can reduce the confounding effects of environmental factors and avoid reverse causation bias [15, 16].

Previous MR studies have found no association between herpes simplex virus (HSV) infection and MS [17], whereas varicella infection is associated with MS [18]. Compared to viral infections, antibody immune responses may not only serve as markers of infection exposure, but also potentially provide clues to the pathophysiological mechanisms associated with ANDs. However, whether there is a causal relationship between these common herpesvirus-induced antibody immune responses and ANDs is unknown, and to the best of our knowledge, our study will be the first to investigate the causal relationship between herpesvirus antibodies and ANDs, thereby further exploring new potential therapeutic targets and potentially informing vaccine selection for patients with ANDs.

## Materials and Methods

### Study Design

MR analysis is a statistical technique used to identify causal relationships, particularly within observational data. It uses genetic variants that influence exposure as instrumental variables (IVs) to assess the causal relationship between exposure and outcomes. A key advantage of MR analysis is its ability to mitigate bias arising from confounding and reverse causation, as genetic variations are generally not influenced by environmental or behavioural factors [15, 16]. MR studies will help to overcome current obstacles in investigating the relationship between antibody immune responses following herpesvirus infection and various ANDs, thus facilitating a more accurate investigation of direct causal relationships.

### Data Source

Our study investigated the immune response of 22 antibodies to five types of herpesvirus infection: Anti-CMV IgG seropositivity, CMV pp28 antibody levels, CMV pp52 antibody levels, CMV pp150 antibody levels; Anti-EBV IgG seropositivity, EBV early antigen-D (EA-D) antibody levels, EBV encoded nuclear antigen-1 (EBNA-1) antibody levels, EBV viral capsid antigen (VCA) p18 antibody levels, EBV ZEBRA antibody levels; Anti-HHV6 IgG seropositivity, Anti-HHV6 IE1A IgG seropositivity, HHV6 IE1A antibody level, Anti-HHV6 IE1B IgG seropositivity, HHV6 IE1B antibody level, HHV6 p101k antibody level, Anti-HHV7 IgG seropositivity, HHV7 U14 antibody levels, Anti-HSV1 IgG seropositivity, HSV1 mgG-1 antibody levels, Anti-HSV2 IgG seropositivity, HSV2 mgG-1 antibody levels; Varicella zoster virus glycoprotein E and I (VZV gE and gI) antibody levels.

The above immune responses were investigated in a Genome-Wide Association Study (GWAS) involving a total of 8,735 individuals for case-control phenotypes and an average (range) of 4,286 (276-8,555) samples per quantitative analysis [19].

Genome-wide summary data for MS consisting of 47,429 cases and 68,374 controls from the European population were obtained from the International MS Genetics Consortium (IMSGC) [20]. To further validate the positive results, UK Biobank (1679 cases and 461254 controls, https://gwas.mrcieu.ac.uk/datasets/ukb-b-17670/) and FinnGen (2620 cases and 449770 controls, https://r11.finngen.fi/pheno/G6_MS) for MS were used.

NMO-related GWAS data were obtained from the largest and most recent NMO GWAS meta-analysis, which included 215 NMO cases and 1,244 controls [21].

The largest MG GWAS reported by Chia et al. was used for the primary analysis in this study, which included 1,873 cases and 36,370 controls [22] (https://www.ebi.ac.uk/gwas/).The MG GWAS focused only on cases positive for anti-acetylcholine receptor antibodies (anti-AChR), excluding individuals positive for antibodies to muscle-specific kinase (anti-MuSK). For external validation, summary statistics were obtained from the UK Biobank (224 cases and 417332 controls, http://www.nealelab.is/uk-biobank) and the FinnGen (504 cases and 449570 controls, https://r11.finngen.fi/pheno/G6_MYASTHENIA).

For GBS, data were obtained from the FinnGen Biobank with 489 cases and 445865 controls (https://r11.finngen.fi/pheno/G6_GUILBAR). Genetic associations for CIDP were derived from 456,348 individuals of European ancestry from the UK Biobank using a fast genome-wide association generalised linear mixed model and were obtained from the catalogue [23]. Detailed information of Data used in our study was shown in supplementary table 1.

### Instrumental Variable Selection

We screened single nucleotide polymorphisms (SNPs) that showed a significant association with exposure factors in the exposure GWAS (P < 5 × 10-8).When the number of SNPs was less than 10, we relaxed the SNPs selection criterion to a p-value threshold of p<1e-07 or p<1e-06, with a minimum standard of p<1e-05 [24]. SNPs with minor allele frequencies (MAF < 0.01) were also deleted, and linkage disequilibrium (LD) between SNPs was resolved by clustering (R2<0.001 within a 10 Mkb clustering distance). Palindromic SNPs were removed. The exposure and outcome data sets were then harmonised. The proportion of variance explained by instrumental variables was calculated using the formula: R2 = 2 ×β2 × EAF × (1 - EAF)/(2 × β2 × EAF × (1 - EAF) + 2 × SE2 × N × EAF × (1 - EAF)), where N is the sample size [25]. The power of the genetic instrument was assessed using the F-statistic, calculated as F= R2 × (N - 2)/(1 - R2). An F-statistic greater than 10 indicates a strong instrument, IVs with F-statistics less than 10 were considered weak and thus excluded from the analysis. The SNiPA(https://snipa.org/snipa3/) and GWAS catalogue(https://www.ebi.ac.uk/gwas/home) was used to check that the SNPs were not associated with other relevant confounders, such as smoking, body mass index (BMI), physical activity, serum 25-hydroxyvitamin D levels.

### Mendelian Randomization Analysis

When there was only one IV, the Wald ratio method was utilized. In cases where two IVs were present, Inverse Variance Weighted (IVW) method will be used. With more than two instrumental variables (IVs) available, univariable Two-Sample MR (TSMR) was used to estimate the total causal effects of antibody immune response on ANDs, we applied five MR analysis methods: IVW, MR Egger, Weighted Median, Simple Mode, and Weighted Mode, IVW served as the primary approach for result interpretation [26].

To clarify whether there were reverse causal relationships between ANDs and antibody immune responses, we also conducted reverse Mendelian randomization.

To account for multiple comparisons, the Bonferroni correction was utilized. We set a significance threshold of α = 0.05/n (where n represents the number of comparisons) to mitigate the increased risk of false positives from multiple testing [27]. This correction method helps maintain the overall Type I error rate at an acceptable level while controlling for the familywise error rate. Therefore, a P-value below 4.55E-04 (calculated as 0.05 divided by the product of 22 comparisons and 5 outcomes) was considered statistically significant.

MR Egger intercept test and Outlier (MR-PRESSO) were used to detect the presence of pleiotropy, When the P-values for MR Egger intercept and MR-PRESSO are less than or equal to 0.05, it suggests the presence of pleiotropic IVs. To address this, we iteratively performed MR Egger intercept and MR-PRESSO tests, removing any outliers each time, until no horizontal pleiotropy was detected. Furthermore, we also employed visual inspection of the scatter plot and forest plot to test the MR “no horizontal pleiotropy”. Additionally, Cochran’s Q statistic was used to assess the degree of heterogeneity among all IVs. P-value from the Q-test greater than 0.05 indicates that there is no significant heterogeneity among the IVs.

To assess the robustness of the identified causal associations, we conducted two sensitivity analyses: the MR-Egger intercept test and leave-one-out analysis. The intercept from the MR-Egger test estimates the degree of directional pleiotropy, while the leave-one-out analysis determines if the significant results are influenced by any single SNP.

### Meta-analysis

For MS and MG, when the MR results of the discovery and replication phases were inconsistent, the meta-analysis was performed to assess combined causality. The choice of effect model was based on the heterogeneity of the results. If there was little significant heterogeneity with I2 ≤ 50%, the fixed-effects model was used to combine the results. Otherwise, the random effects model was used.

The statistical analyses in this study were performed using R version 4.4.1, including TwoSampleMR, MRPRESSO and Metafor. The study protocol and details were not pre-registered

## Results

### Selection of Instrumental Variables

Detailed information on the IVs of antibody immune responses to five types of herpesvirus is shown in Supplementary Table 2. The P value thresholds for anti-CMV IgG seropositivity, CMV pp28 antibody levels, CMV pp52 antibody levels, CMV pp150 antibody levels, anti-EBV IgG seropositivity, anti-HHV6 IgG seropositivity, anti-HHV6 IE1A IgG seropositivity, HHV6 IE1A antibody levels, anti-HHV6 IE1B IgG seropositivity, HHV6 IE1B antibody levels, HPV6 p101k antibody levels, anti-HHV7 IgG seropositivity, anti-HSV 1 IgG seropositivity, HSV1 mgG-1 antibody levels anti-HSV2 IgG seropositivity and HSV2 mgG-1 antibody levels were 1e-05, HHV7 U14 antibody levels were 1e-07, EBV EA-D antibody levels, EBV EBNA-1 antibody levels, EBV VCA p18 antibody levels, EBV ZEBRA antibody levels and VZV gE and gI antibody levels were 5e-08. rs1808192 in HHV7 U14 antibody levels, rs34811474 in anti-HSV 1 IgG seropositivity and rs13204572 in VZV gE and gI antibody levels were deleted as they were associated with BMI. IVs for MS, NMO, MG, CIDP, GBS are shown in Supplementary Tables 3, 5, 7, 9, 11, respectively. All F values of the IVs were greater than 10, indicating that weak instruments were excluded.

### Antibody Immune Response Following Herpesvirus Infection on MS

In this study, the antibody unit was the logarithmic transformation of the antibody median fluorescence intensity. We observed a causal effect of a 1-unit increase in EBV EBNA-1 antibody levels, EBV VCA p18 antibody levels and HHV7 U14 antibody levels correlating with an increasing risk of MS [IVW: odds ratio (OR) = 3. 009, 95% CI: 2.377-3.807, P=4.772e-20, adjusted p=2.250e-18; IVW: OR=8.700, 95% CI: 6.128-12.351, P=1.059e-33, adjusted p=1.165e-31 and IVW: OR=3.359, 95%CI: 2.034-5.546, P=2.187e-06, adjusted p=2.406e-04]. In addition, causal evidence was also found for a differential effect of a 1-unit increase in EBV ZEBRA antibody levels (IVW: OR = 0.592, 95% CI: 0.491-0.713, P = 3.195e-08, adjusted p =3.515e-6), the analysis did not reveal causal relationships between other antibodies and the risk of MS (Fig 1, S Table 4). To validate the above results, we also used the UK Biobank and FinnGen MS datasets, with detailed results shown in Table 1. In all three datasets, the causal effects of EBV EBNA antibody levels and EBV VCA p18 antibody levels on MS were significant; however, the MR results of HHV7 U14 antibody levels and EBV ZEBRA antibody levels on MS differed between the three datasets, and the meta-analysis did not confirm that the overall effects of these on MS were significant.

**Fig 1:**
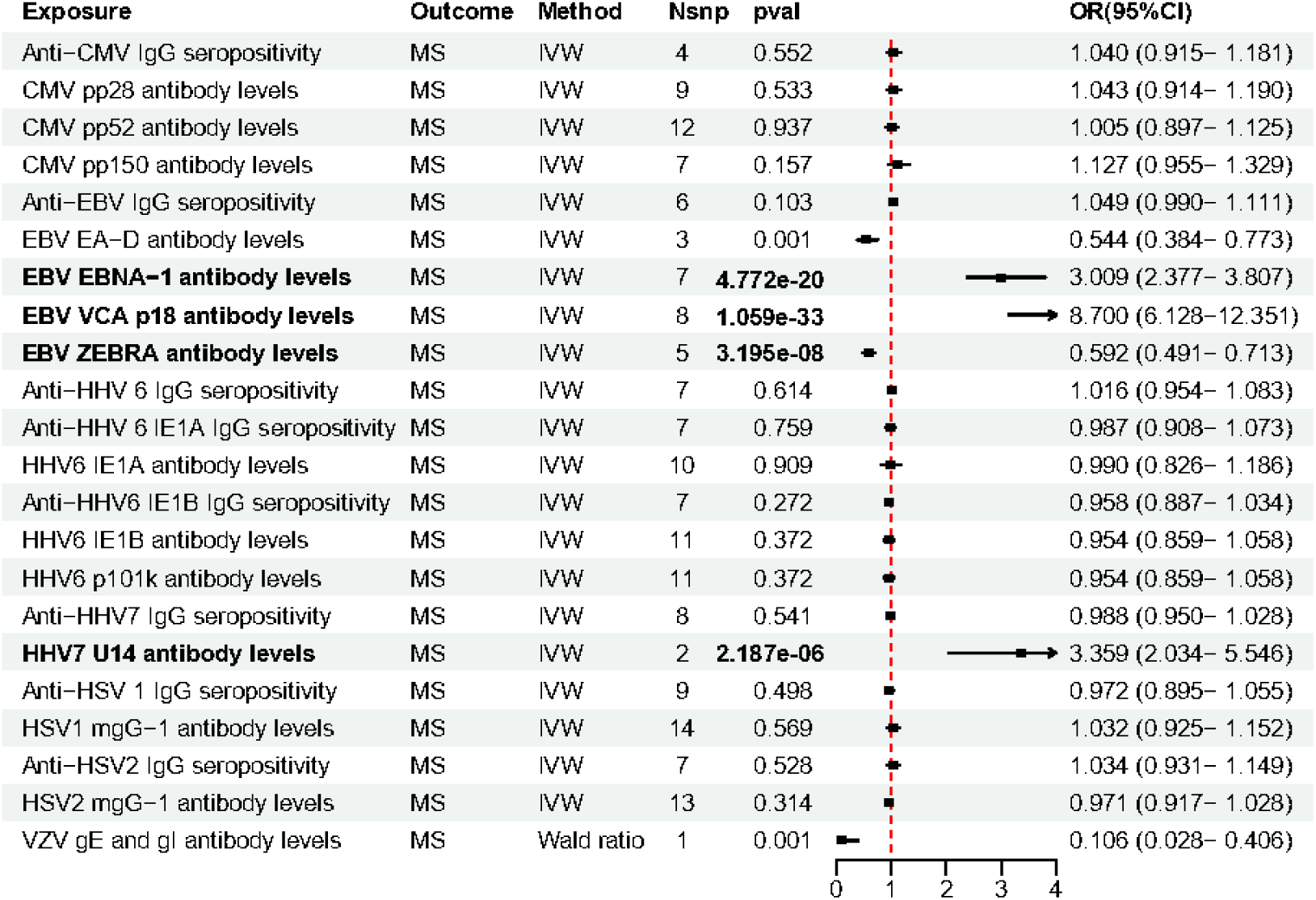
MR estimate results of antibodies of five herpesviridae viruses on MS.

**Table 1.**
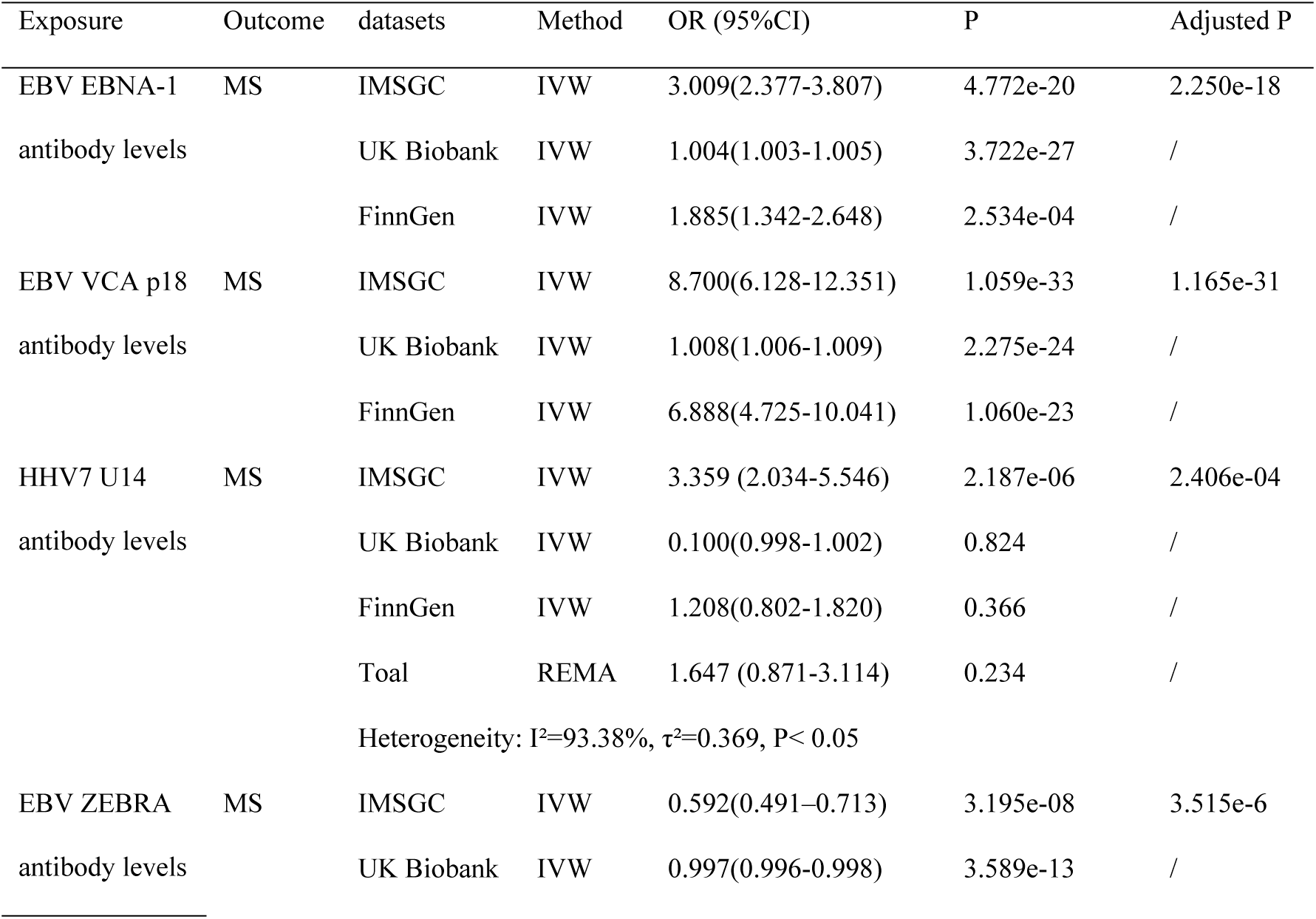
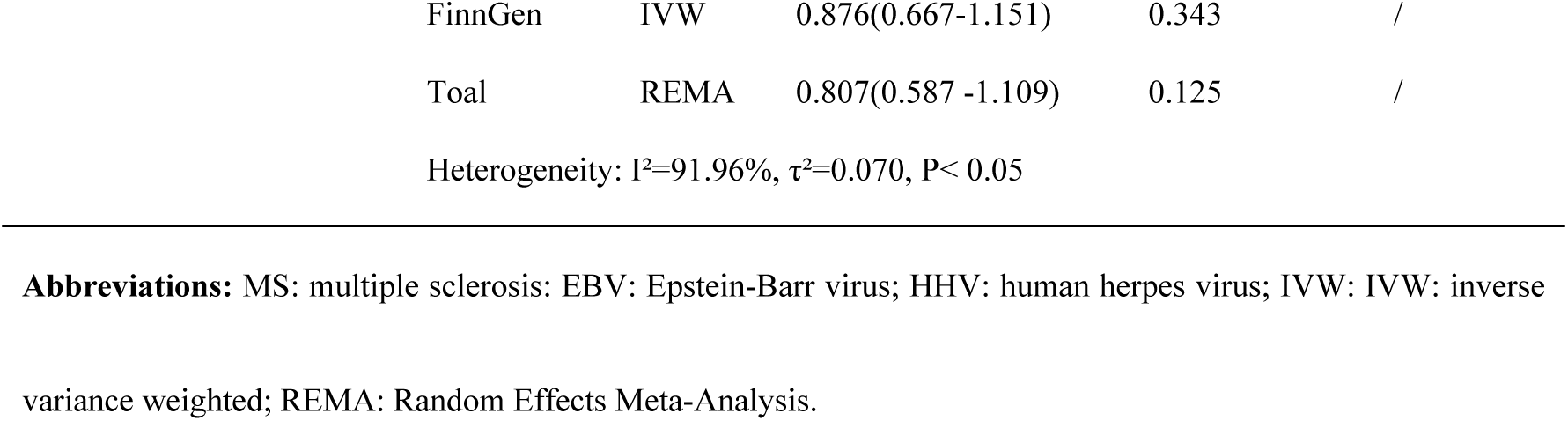
Mendelian Randomization Analysis of the Causal Effects of Herpesvirus Antibodies on Multiple Sclerosis Risk Across Different Datasets.

### Pleiotropy and Sensitivity Analysis for MS

Heterogeneity was observed among the IVs of EBV EBNA-1 antibody levels (Q=27.055 P=1.414e-4), EBV VCA p18 antibody levels (Q=70.545 P=1.147e-12), HHV6 IE1A antibody levels (Q=17.036 P=0.048) and anti-HSV2 IgG seropositivity (Q=13.885 P=0.031) in terms of their effect on MS (S Table 4). All p-values from the MR Egger intercepts and MR-PRESSO global tests were greater than 0.05, indicating the absence of outlier SNPs and no evidence of horizontal pleiotropy between the IVs and outcomes. Leave-one-out analysis showed that the results remained consistent regardless of which individual SNP was excluded (S Fig 1-22).

It is noteworthy that the final IVs of HHV7 U14 antibody levels and Varicella zoster virus glycoproteins E and I (VZV gE and gI) antibody levels, filtered by the above steps, consist of only two SNPs. IVs of the former have no heterogeneity (Q=2.823 P=0.093), single SNP MR analysis showed the similar result (rs7773105: P=2.175e-05, adjusted P=0.002, OR=2.562, 95%CI:1.660-3.956; rs805282: P=3.911e-12, adjusted P=4.300e-11, OR=4.277, 95%CI: 2.97-6.448), we consider the causal relationship between HHV7 U14 antibody levels and MS to be reliable. However, there was significant heterogeneity between these two IVs of VZV gE and gI antibody levels (Q= 53.561 P= 2.507e-13). In the single SNP MR analysis, the significance of the effects of these two IVs on MS varied (rs9270551: P = 2.78E-276, adjusted P = 3.058e-274, OR = 0.557e-06, 95% CI: 0.369e-03 - 0.843e-03; rs9272285: P = 1.052E-03, adjusted P = 0.116, OR = 0.106, 95% CI: 0.028-0.406). In particular, rs9270551 was strongly associated with MS (P = 1E-200), suggesting the presence of horizontal pleiotropy. Therefore, based on the MISGC databases, we used rs9272285 as an IV for VZV gE and gI antibody levels for MS. To further investigate the association between VZV gE and gI antibody levels and MS, we also performed MR analysis based on the MS datasets in UK Biobank and FinnGen with the following results: OR (95%CI) =1.000(0.999-1.001), P=0.299 (UK Biobank), 0.2171830962485 OR (95%CI) =0.820 (0.5978-1.124) P=0.217 (FinnGen), so there wasn’t the causal relationship between VZV gE and gI antibody levels and MS.

### MS on Antibody Immune Response

Reverse Mendelian randomization revealed that MS does not have significant reverse causal effects on antibodies to the aforementioned herpesvirus antigens, with all p-values greater than the adjusted p-value (4.550e-04) (S Fig 23).

### Antibody Immune Response on NMO

There is a causal relationship between elevated levels of two EBV-related antibodies and a reduced risk of NMO. These two antibodies are EBV EA-D antibody levels (IVW: OR = 0.043 95% CI: 0.010-0.196, P = 4.546e-05, adjusted p=0.005) and EBV ZEBRA antibody levels (IVW: OR = 0.079, 95% CI: 0.043-0.147, P = 7.587e-16, adjusted p=8. 345e-14). Conversely, an increased risk of NMO could be caused by higher antibody levels of HHV7 U14 (IVW: OR =10.641 95% CI: 3.025-37.429, P=2.285e-04 adjusted p=0.025) or VZV gE and gI (IVW: OR =17.220 95% CI: 9.760-30.380, P=8.727e-23 adjusted p=9.599e-21). Antibodies to other viruses of the Herpesviridae family had no causal effect on the risk of NMO (Fig 2, S Table 6).

**Fig 2.**
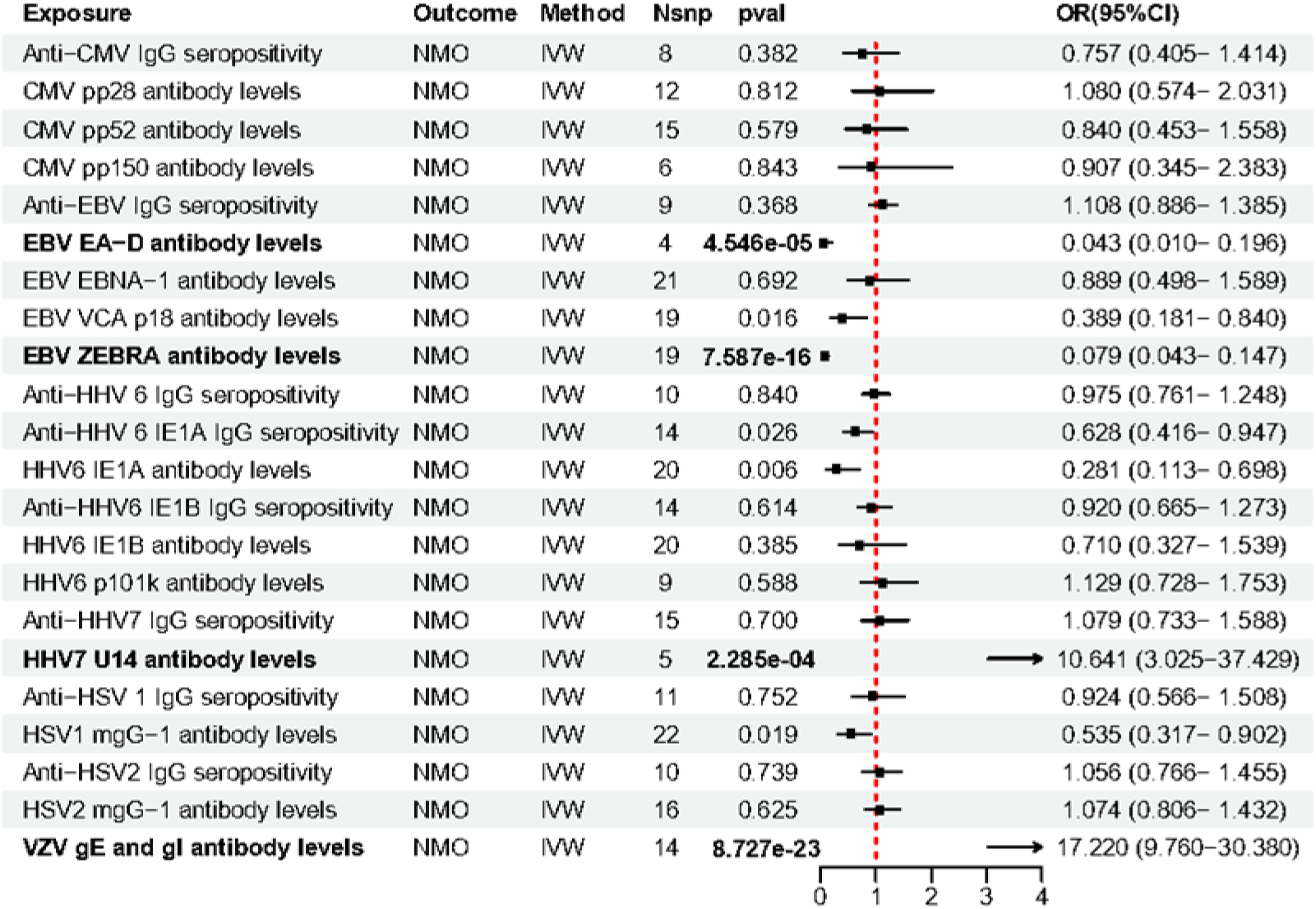
MR estimate results of antibodies of five herpesviridae viruses on NMO.

### Pleiotropy and Sensitivity Analysis for NMO

Heterogeneity was observed in the IVs for EBV EA-D antibody levels (Q=6.892, P=0.075), EBV EBNA-1 antibody levels (Q=70.545, P=6.776e-4), EBV ZEBRA antibody levels (Q=34. 471, P=0.011), HHV6 IE1A antibody levels (Q=29.584, P=0.042) and HHV6 IE1B antibody levels (Q=31.661, P=0.034) on NMO (Supplementary Table 6). The MR-PRESSO global test indicated the presence of horizontal pleiotropy between the IVs for EBV EBNA-1 antibody levels on NMO (P=0.001), EBV ZEBRA antibody levels (P=0.022) and HHV6 IE1B antibody levels (P=0.034). However, the p-values of the Egger intercept were greater than 0.05 and no significant outliers were detected. The leave-one-out analysis confirmed that no single SNP significantly altered the overall results. Therefore, the results still have some informative value.

For the IVs of EBV ZEBRA antibody levels, the p-value of the MR-PRESSO global test was less than 0.05, but the Egger intercept was greater than 0.05. All five MR statistical methods showed a consistent direction of effect, similar effect estimates and OR values (Supplementary Table 6). The leave-one-out analysis again showed that the results were robust to the exclusion of any single SNP (Supplementary Fig 9). Thus, the overall results continue to support the existence of a causal relationship between EBV ZEBRA antibody levels and NMO.

Pleiotropy tests for the IVs of other antibodies showed no outlier SNPs and no evidence of horizontal pleiotropy between them and the outcomes (Supplementary Table 6). Leave-one-out analysis consistently showed that excluding any single SNP did not affect the results (S Fig 1-22).

### NMO on Antibody Immune Response

Reverse causal effects of NMO on antibodies were not found (S Fig 24).

### Antibody Immune Response on MG

We found a causal effect between increased VZV gE and gI antibody levels and an increased risk of MG (IVW: OR = 2.542 95% CI: 1.972-3.277, P =5.897e-13 adjusted p= 2.594e-12). No significant associations were found between other antibodies and MG (Fig 3). In addition, the UK Biobank and FinnGen MG datasets were used to validate this result, with detailed results shown in Table 2. In all three datasets, the causal effect of VZV gE and gI antibody levels on MG was all significant.

**Fig 3.**
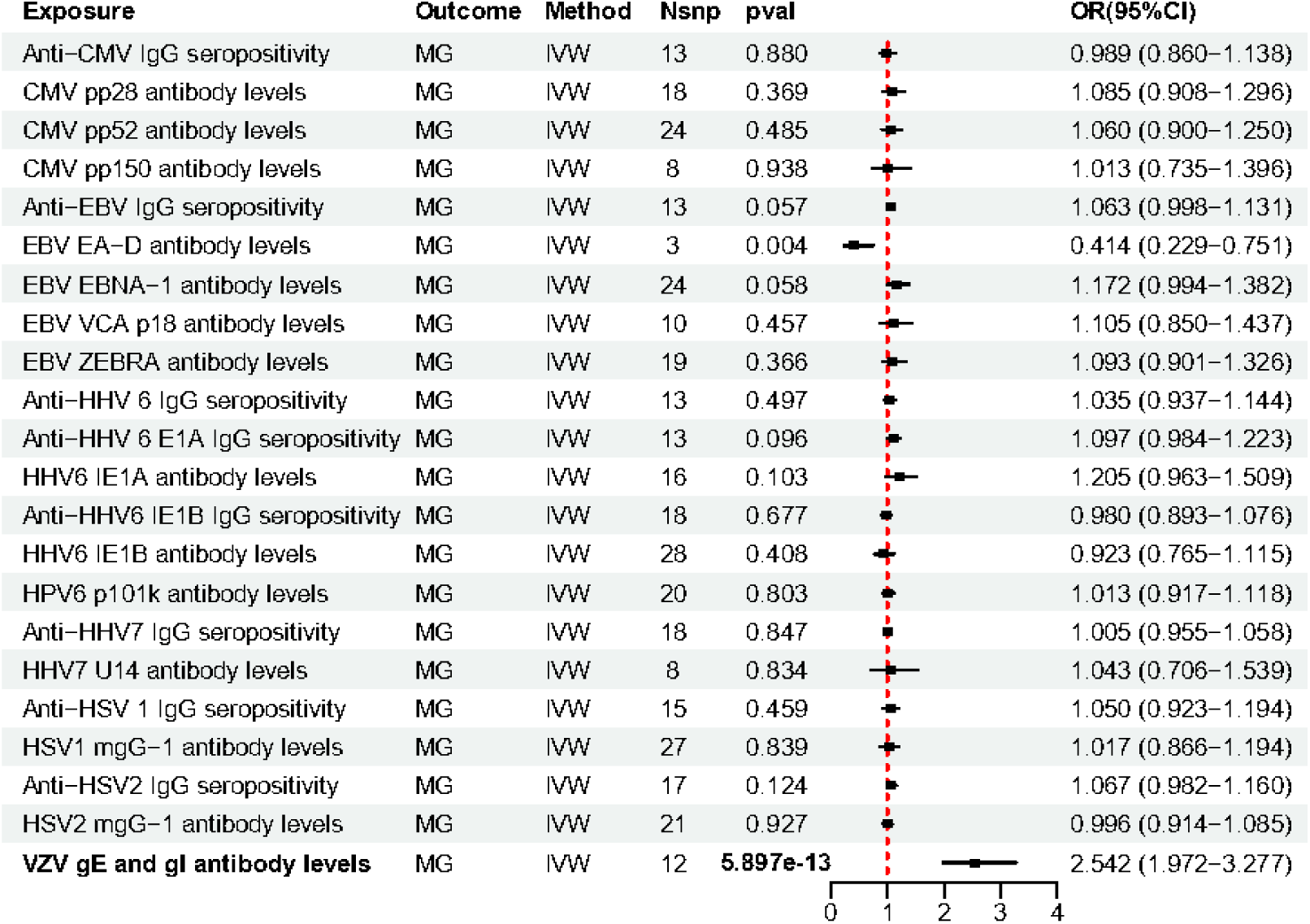
MR estimate results of antibodies of five herpesviridae viruses on MG.

**Table 2.** MR estimate results of antibodies of VZV gE and gI antibody levels on MG.

| Exposure | Outcome | Author and datasets | Method | OR (95%CI) | P | Adjusted P |
| --- | --- | --- | --- | --- | --- | --- |
| VZV gE and gI antibody levels | MG | Jiang L et al. | IVW | 2.542(1.972-3.277) | 5.897e-13 | 2.594e-12 |
|  |  | UK Biobank | IVW | 1.00024(1.000035-1.000445) | 0.022 | / |
|  |  | FinnGen | IVW | 2.619(1.768-3.880) | 1.576e-06 | / |

**Abbreviations:** MG: myasthenia gravis; VZV: varicella zoster virus; IVW: inverse variance weighted; REMA: random effects meta-Analysis.

### Pleiotropy and Sensitivity Analysis for MG

IVs of EBV EA-D antibody levels (Q=44.715, P=0.004) and EBV ZEBRA antibody levels (Q=37.615, P=0.004) show significant heterogeneity (Supplementary Table 8). Although MR-PRESSO identified pleiotropy, the Egger intercept did not indicate significant pleiotropy and no significant outliers were detected. IVs from other antibodies did not show heterogeneity and pleiotropy (S Table 8). The leave-one-out analysis gave similar results, even after excluding individual SNPs (S Fig 1-22).

### MG on Antibody Immune Responses

The adjusted P values from the MR analysis indicated that there was no causal relationship between MG and any of the 22 types of antibody immune response (S Fig 24).

### Causal Relationships Detected Between Antibody Immune Responses and GBS/CIDP

We found no forward or reverse causal relationships between the 22 antibody immune responses to five types of herpesviruses and GBS and CIDP (Fig 4-5; S Fig 26-27). IVs for HSV1 mgG-1 antibody levels on CIDP showed heterogeneity (Q = 44.905, P = 0.03) (S Table 12). Pleiotropy was detected by MR-PRESSO but not by the Egger intercept, and the MR-PRESSO global test did not identify any significant outliers. Leave-one-out analysis showed no significant differences in MR results after removing individual SNPs (S Fig 1-22).

**Fig 4.**
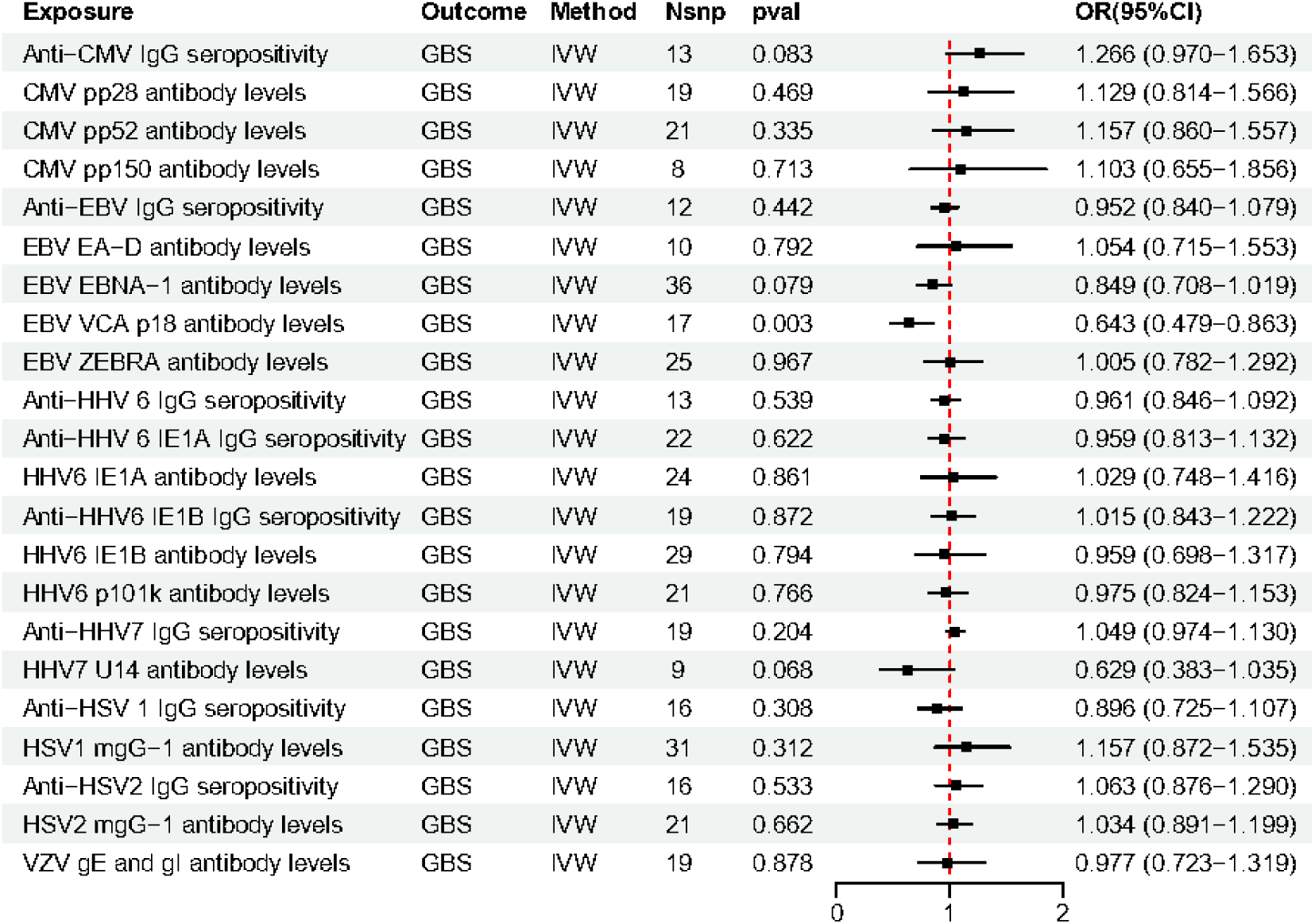
MR estimate results of antibodies of five herpesviridae viruses on GBS.

**Fig 5.**
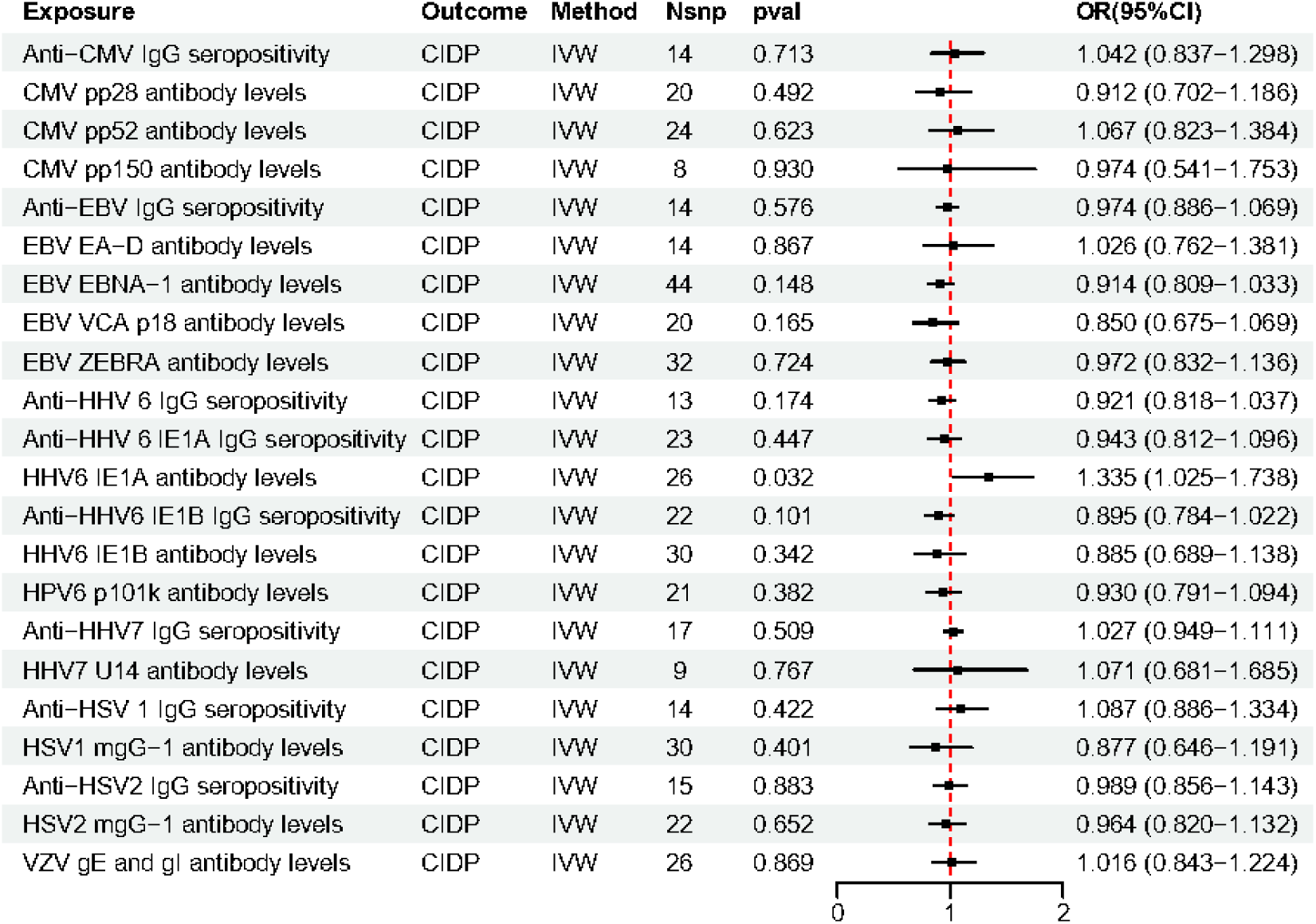
MR estimate results of antibodies of five herpesviridae viruses on CIDP.

All other results, including those for GBS and CIDP, passed the heterogeneity and pleiotropy tests. The sensitivity analysis did not identify any SNPs that significantly affected the overall results.

## Discussion

As the MR results showed, EBV EBNA-1 antibody and VCA p18 antibody were positively correlated with MS. Levels of EBV ZEBRA antibody were negatively correlated with MS in the IMSGC and UK Biobank datasets, but not in FinnGen, and the meta-analyses did not confirm a causal relationship. In addition, HHV7 U14 antibody levels were positively associated with MS in the IMSGC dataset, but not in the UK Biobank or FinnGen datasets, and the overall causal relationship was not established by the meta-analyses.

Negative associations between EBV ZEBRA and EA-D antibody levels and NMO were shown in this study. Conversely, a positive association was found between NMO and antibody levels of VZV gE, VZV gI and HHV7 U14. VZV gE and gI antibody levels showed a positive association with MG.

The above results suggest an important role for EBV antigen-associated antibodies in the pathogenesis of AND. EBV infects and persists in human B cells by exploiting the B cell environment to sustain its life cycle while evading the host immune system through minimal expression of viral proteins [28]. Several antibodies to EBV proteins can be detected in EBV carriers. These proteins include EBNA-1 (BKRF1), a latency protein; VCA p18 (BFRF3), a structural protein of the mature virus; ZEBRA (BZLF1), a lytic cycle protein; and EA-D, a basic component of the viral DNA replication complex. [28, 29]. Previous studies have confirmed that higher antibody levels against EBNA-1 and VCA p18 (or VCA) are associated with an increased risk of MS; in individuals with genetic factors associated with lower EBNA-1 IgG titres, the onset of MS was delayed by 3.5 years [30, 31], [32]. A region within the carboxyl half of EBNA1 (amino acids 385-420) induces elevated antibody levels in MS patients and shows the strongest association with MS risk compared to antibody levels against other domains of EBNA1 [33]. EBNA1-specific antibodies are found in the cerebrospinal fluid (CSF) of some MS patients; these antibodies may play a role in the formation of oligoclonal bands, which are antibodies produced by clonal B-cell-derived plasma cells in the central nervous system and are a hallmark of MS [34]. Injection of a peptide derived from the aa395-420 region of EBNA1 into mice with experimental autoimmune encephalomyelitis (EAE) exacerbated CNS autoimmunity [34, 35].

Our research confirms a causal relationship between antibodies to EBNA-1 or VCA p18 and MS. However, the exact mechanisms by which these two antibodies cause MS are not yet clear. Based on the current evidence, molecular mimicry of EBNA-1 and VCA p18 may play some role.Antibodies against VCA p18 (BFRF3) cross-react with the cytoplasmic protein septin-9 [36]. Septin-9, a cytoplasmic protein that forms filaments, may play a role in cytokinesis, exocytosis, neurite outgrowth and cell cycle control. While it is widely expressed in various tissues, it is emerging as an intriguing candidate autoantigen for MS due to gene mutations that cause hereditary neuralgic amyotrophy [37]. In the CNS, septin-9 is mainly found in neurons and its expression is upregulated after trauma [38]. Antibodies to EBNA1 have been observed to cross-react with anoctamin 2, ɑ B-crystallin, myelin basic protein (MBP) and glial cell adhesion molecule [33, 34, 39]. However, the pathogenic role of these antibodies remains unclear because successful cell-depleting therapy for MS does not affect antibody-producing plasma cells in the short term. Moreover, such treatment does not seem to reduce oligoclonal bands [7]. These two antibodies may become new therapeutic targets in MS, particularly in patient groups with high levels of these antibodies and poor response to other treatments.

ZEBRA (BZLF1) was considered to be a major environmental factor associated with MS and a significant contributor to the disease process [40]. BZLF1 was found in 46% of MS brains, mainly associated with chronic lesions, and in 44% of non-MS brain tissue [41]. ZEBRA is recognised as one of the viral immediate early genes that play an important role in the transition of EBV from latency to lytic replication [42], Reactivation of latent EBV infection is associated with disease activity in MS patients; however, research results are controversial [43–45]. Meanwhile, increasing EBV ZEBRA antibodies have not been found in MS [46], the role of ZEBRA antibodies in MS remains unclear.

Previous research found that anti-EA seropositivity was significantly higher in the NMOSD group compared to the HC group and MS patients [47, 48], but another study suggested that exposure to Epstein-Barr virus was not associated with an increased likelihood of NMO compared to healthy subjects [49]. Although our research suggests that BV ZEBRA and EA-D antibody levels may be associated with a reduced risk of NMO, further studies are needed to confirm this. Based on the advantages of MR statistics, this study suggests that EBV antibodies may provide new clues for NMO treatment. In particular, for NMO patients with high levels of EBV antibodies, understanding the different roles of different antibodies in the pathogenesis of NMO may facilitate the exploration of personalised treatment approaches.

Current research on the association between HHV infection and the risk of MS has yielded controversial results, [50–52], This controversy is reflected in the results of this MR analysis. A meta-analysis found that CMV IgG seroprevalence was lower in people with MS than in controls in Europe [53]. However, our study does not support a causal relationship between CMV antibody immune response and risk of MS. Although one MR study suggested a causal relationship between herpesvirus infection and MS, we did not find a causal relationship between VZV gE and gI antibody levels and MS [18].

At present, the relationship between HHV and NMO is not very clear. Our study suggests that HHV may be one of the important pathogens of NMO, and HHV 7 U14 antibody may be detrimental to NMO, but its exact pathogenic role in NMO requires further investigation. VZV has been considered as a precursor infectious agent of NMO [54–56], Causal association between chickenpox infection and NMO was also recently confirmed by MR study [57], Our study further confirmed a causal effect of VZV gE and gI antibody levels on NMO. VZV infection is an important risk factor for NMO, and vaccination against VZV may help to reduce the risk of NMO. For NMO patients with high levels of VZV gE and gI antibodies, the immune response and related pathogenic mechanisms induced by VZV infection could potentially become a new therapeutic approach.

Among 22 types of antibody immune response, only VZV gE and gI antibody levels were found to be associated with MG. The resultant GWAS data from two different datasets both confirm this causal association, suggesting that unlike in MS, the pathogenic role of VZV in MG was robust, which may imply new therapeutic targets for MG, especially for patients with high levels of this antibody.

This study has a number of important strengths. First, we effectively reduced the influence of confounding factors and improved the accuracy and reliability of our results by using a novel MR analysis method. Second, our comprehensive analysis explored the causal relationships between 22 antibody immune responses to five herpesvirus infections and five types of ANDs. This provided a detailed understanding of their interactions. Third, we applied a rigorous correction procedure to ensure the robustness and validity of our findings. For positive results, we cross-validated with additional publicly available GWAS outcome data. We also conducted further meta-analyses to resolve any inconsistencies. These measures helped to reduce the risk of false positives. They also strengthened the credibility of our conclusions.

Although this study provides valuable insights, it has several limitations that should be acknowledged. First, the use of summary level data in MR analyses, although powerful, may introduce bias due to the assumption of linearity and the inability to explore non-linear relationships. In addition, the generalisability of the findings is limited by the demographic and genetic composition of the datasets used, which are predominantly of European ancestry. This raises questions about the applicability of the results to more diverse populations. Another limitation is the potential for pleiotropy, where genetic variants used as instruments may affect the outcome through pathways other than the exposure of interest, leading to biased estimates. Although we used methods to mitigate pleiotropy, such as MR-PRESSO, the possibility of residual confounding cannot be completely excluded. Finally, the study relies on the availability and quality of existing GWAS datasets, which may vary in their coverage and accuracy of antibody measurements, potentially affecting the robustness of the findings. Future studies should aim to validate these findings in more diverse populations and with individual-level data to better understand the complex relationships between herpesvirus antibodies and autoimmune neurodegenerative diseases.

## Conlusion

The results of this study reveal predicted causal effects of EBV EBNA-1 antibody, EBV VCA p18 antibody on MS, HHV7 U14, VZV gE and gI antibody, EBV EA-D antibody and EBV ZEBRA antibody on NMO, VZV gE and gI antibody on MG, This might suggest potential role for above antibodies in the pathogenesis of ANDs, also This finding highlights the need for further exploration into the specific mechanisms of above antibodies in the pathophysiological processes of ANDs, providing an important theoretical basis for the development of more effective treatment strategies in the future.

## Declarations

### Ethics approval and consent to participate

Publicly available datasets were used for all data in our study. Informed consent was obtained from all participants prior to enrollment, and ethical approval was obtained for each cohort.

### Consent for Publication

All authors provide their consent for the publication of this manuscript.

### Availability of data and materials

The original papers presented in this study can be found in the supplementary materials.

### Authors’ contributions

Sunjun Sun: Writing – original draft, Visualization, Validation, Methodology, Formal analysis, Funding acquisition;

Yiyong Wen: Writing-original draft, Formal analysis, review & editing;

Pan Tang: Writing– review & editing, Validation.

Fan Xie: Writing – review & editing, Visualization, Validation.

Jun Wen: Writing – review & editing, Supervision,

Anding Xu: Writing –Conceptualization, review & editing, Supervision, Funding acquisition. Project administration.

## Data Availability

The original papers presented in this study can be found in the supplementary materials.

## Acknowledgements

The authors thank the Multiple Sclerosis Genetics Consortium and the International Myasthenia Gravis Genomics Consortium, the UK Biobank and the FinnGen study for providing publicly available MS, MG and GBS GWAS summary statistics for this analysis, as well as the studies by Butler-Laporte G et al, Estrada K et al, Chia R et al, Jiang L et al for antibody immune responses to five types of herpesvirus, NMO, MG, CIDP GWAS summary statistics.

## List of abbreviations

ANDs: Autoimmune neuroinflammatory diseases
MS: multiple sclerosis
NMO: neuromyelitis optica
GBS: Guillain-Barr é syndrome
CIDP: chronic inflammatory demyelinating polyneuropathy
MG: myasthenia gravis
CNS: central nervous system
PNS: peripheral nervous system
NJM: neuromuscular junction
EBV: Epstein-Barr virus
HHV 6A: Human herpesvirus 6A;
CMV: Cytomegalovirus
AQP4 IgG+ NMOSD: aquaporin-4 antibody-positive NMO spectrum disord
MR: mendelian randomisation
HSV: herpes simplex virus
EBNA-1: EBV encoded nuclear antigen-1
EBV-VCA p18: EBV viral capsid antigen p18
VZV gE and gI: Varicella zoster virus glycoprotein E and I
GWAS: Genome-Wide Association Study
IMSGC: International MS Genetics Consortium
anti-AChR: anti-acetylcholine receptor antibodies
anti-MuSK: antibodies to muscle-specific kinase
SNPs: single nucleotide polymorphisms
MAF: minor allele frequencies
LD: linkage disequilibrium
BMI: body mass index
IVW: Inverse Variance Weighte
IVs: instrumental variables (IVs)
TSMR: Two-Sample MR
MR-PRESSO: MR Egger intercept test and Outlier
OR: odds ratio
CSF: cerebrospinal fluid
EAE: experimental autoimmune encephalomyelitis.

## References

1. Khan AW, Farooq M, Hwang MJ, Haseeb M, Choi S. Autoimmune Neuroinflammatory Diseases: Role of Interleukins. Int J Mol Sci. 2023;24(9). Epub 20230427. doi: 10.3390/ijms24097960. PubMed PMID: 37175665; PubMed Central PMCID: PMCPMC10178921.

2. Bhagavati S. Autoimmune Disorders of the Nervous System: Pathophysiology, Clinical Features, and Therapy. Front Neurol. 2021;12:664664. Epub 20210414. doi: 10.3389/fneur.2021.664664. PubMed PMID: 33935958; PubMed Central PMCID: PMCPMC8079742.

3. Zarobkiewicz MK, Morawska I, Michalski A, Roliński J, Bojarska-Junak A. NKT and NKT-like Cells in Autoimmune Neuroinflammatory Diseases-Multiple Sclerosis, Myasthenia Gravis and Guillain-Barre Syndrome. Int J Mol Sci. 2021;22(17). Epub 20210901. doi: 10.3390/ijms22179520. PubMed PMID: 34502425; PubMed Central PMCID: PMCPMC8431671.

4. Gogia B, Rocha Cabrero F, Khan Suheb MZ, Lui F, Rai PK. Chronic Inflammatory Demyelinating Polyradiculoneuropathy. StatPearls. Treasure Island (FL): StatPearls Publishing Copyright © 2024, StatPearls Publishing LLC.; 2024.

5. Miller FW. The increasing prevalence of autoimmunity and autoimmune diseases: an urgent call to action for improved understanding, diagnosis, treatment, and prevention. Curr Opin Immunol. 2023;80:102266. Epub 20221126. doi: 10.1016/j.coi.2022.102266. PubMed PMID: 36446151; PubMed Central PMCID: PMCPMC9918670.

6. Pisetsky DS. Pathogenesis of autoimmune disease. Nat Rev Nephrol. 2023;19(8):509–24. Epub 20230510. doi: 10.1038/s41581-023-00720-1. PubMed PMID: 37165096; PubMed Central PMCID: PMCPMC10171171.

7. Bjornevik K, M ü nz C, Cohen JI, Ascherio A. Epstein-Barr virus as a leading cause of multiple sclerosis: mechanisms and implications. Nat Rev Neurol. 2023;19(3):160–71. Epub 20230209. doi: 10.1038/s41582-023-00775-5. PubMed PMID: 36759741.

8. Bjornevik K, Cortese M, Healy BC, Kuhle J, Mina MJ, Leng Y, et al. Longitudinal analysis reveals high prevalence of Epstein-Barr virus associated with multiple sclerosis. Science. 2022;375(6578):296–301. Epub 20220113. doi: 10.1126/science.abj8222. PubMed PMID: 35025605.

9. Miao Y, Shi Z, Zhang W, Zhu L, Tang S, Chen H, et al. Immune Repertoire Profiling Reveals Its Clinical Application Potential and Triggers for Neuromyelitis Optica Spectrum Disorders. Neurol Neuroimmunol Neuroinflamm. 2023;10(5). Epub 20230706. doi: 10.1212/nxi.0000000000200134. PubMed PMID: 37414573; PubMed Central PMCID: PMCPMC10326831.

10. Kuwabara S. Guillain-Barré syndrome: epidemiology, pathophysiology and management. Drugs. 2004;64(6):597–610. doi: 10.2165/00003495-200464060-00003. PubMed PMID: 15018590.

11. Rodríguez Y, Vatti N, Ramírez-Santana C, Chang C, Mancera-Páez O, Gershwin ME, Anaya JM. Chronic inflammatory demyelinating polyneuropathy as an autoimmune disease. J Autoimmun. 2019;102:8–37. Epub 20190506. doi: 10.1016/j.jaut.2019.04.021. PubMed PMID: 31072742.

12. Kakalacheva K, Maurer MA, Tackenberg B, Münz C, Willcox N, Lünemann JD. Intrathymic Epstein-Barr virus infection is not a prominent feature of myasthenia gravis. Ann Neurol. 2011;70(3):508–14. doi: 10.1002/ana.22488. PubMed PMID: 21905082.

13. Cavalcante P, Marcuzzo S, Franzi S, Galbardi B, Maggi L, Motta T, et al. Epstein-Barr virus in tumor-infiltrating B cells of myasthenia gravis thymoma: an innocent bystander or an autoimmunity mediator? Oncotarget. 2017;8(56):95432–49. Epub 20170908. doi: 10.18632/oncotarget.20731. PubMed PMID: 29221139; PubMed Central PMCID: PMCPMC5707033.

14. Lawlor DA, Harbord RM, Sterne JA, Timpson N, Davey Smith G. Mendelian randomization: using genes as instruments for making causal inferences in epidemiology. Stat Med. 2008;27(8):1133–63. doi: 10.1002/sim.3034. PubMed PMID: 17886233.

15. Smith GD, Ebrahim S. ’Mendelian randomization’: can genetic epidemiology contribute to understanding environmental determinants of disease? Int J Epidemiol. 2003;32(1):1–22. doi: 10.1093/ije/dyg070. PubMed PMID: 12689998.

16. Zheng J, Baird D, Borges MC, Bowden J, Hemani G, Haycock P, et al. Recent Developments in Mendelian Randomization Studies. Curr Epidemiol Rep. 2017;4(4):330–45. Epub 20171122. doi: 10.1007/s40471-017-0128-6. PubMed PMID: 29226067; PubMed Central PMCID: PMCPMC5711966.

17. Zhang W, Wu P, Yin R, Sun M, Zhang R, Liao X, et al. Mendelian Randomization Analysis Suggests No Associations of Herpes Simplex Virus Infections With Multiple Sclerosis. Front Neurosci. 2022;16:817067. Epub 20220301. doi: 10.3389/fnins.2022.817067. PubMed PMID: 35299622; PubMed Central PMCID: PMCPMC8920987.

18. Zhu G, Zhou S, Xu Y, Gao R, Zhang M, Zeng Q, et al. Chickenpox and multiple sclerosis: A Mendelian randomization study. J Med Virol. 2023;95(1):e28315. Epub 20221123. doi: 10.1002/jmv.28315. PubMed PMID: 36380510.

19. Butler-Laporte G, Kreuzer D, Nakanishi T, Harroud A, Forgetta V, Richards JB. Genetic Determinants of Antibody-Mediated Immune Responses to Infectious Diseases Agents: A Genome-Wide and HLA Association Study. Open Forum Infect Dis. 2020;7(11):ofaa450. Epub 20200924. doi: 10.1093/ofid/ofaa450. PubMed PMID: 33204752; PubMed Central PMCID: PMCPMC7641500.

20. Multiple sclerosis genomic map implicates peripheral immune cells and microglia in susceptibility. Science. 2019;365(6460). doi: 10.1126/science.aav7188. PubMed PMID: 31604244; PubMed Central PMCID: PMCPMC7241648.

21. Estrada K, Whelan CW, Zhao F, Bronson P, Handsaker RE, Sun C, et al. A whole-genome sequence study identifies genetic risk factors for neuromyelitis optica. Nat Commun. 2018;9(1):1929. Epub 20180516. doi: 10.1038/s41467-018-04332-3. PubMed PMID: 29769526; PubMed Central PMCID: PMCPMC5955905.

22. Chia R, Saez-Atienzar S, Murphy N, Chi ò A, Blauwendraat C, Roda RH, et al. Identification of genetic risk loci and prioritization of genes and pathways for myasthenia gravis: a genome-wide association study. Proc Natl Acad Sci U S A. 2022;119(5). doi: 10.1073/pnas.2108672119. PubMed PMID: 35074870; PubMed Central PMCID: PMCPMC8812681.

23. Jiang L, Zheng Z, Fang H, Yang J. A generalized linear mixed model association tool for biobank-scale data. Nat Genet. 2021;53(11):1616–21. Epub 20211104. doi: 10.1038/s41588-021-00954-4. PubMed PMID: 34737426.

24. Burgess S, Butterworth A, Thompson SG. Mendelian randomization analysis with multiple genetic variants using summarized data. Genet Epidemiol. 2013;37(7):658–65. Epub 20130920. doi: 10.1002/gepi.21758. PubMed PMID: 24114802; PubMed Central PMCID: PMCPMC4377079.

25. Shim H, Chasman DI, Smith JD, Mora S, Ridker PM, Nickerson DA, et al. A multivariate genome-wide association analysis of 10 LDL subfractions, and their response to statin treatment, in 1868 Caucasians. PLoS One. 2015;10(4):e0120758. Epub 20150421. doi: 10.1371/journal.pone.0120758. PubMed PMID: 25898129; PubMed Central PMCID: PMCPMC4405269.

26. Sekula P, Del Greco MF, Pattaro C, Köttgen A. Mendelian Randomization as an Approach to Assess Causality Using Observational Data. J Am Soc Nephrol. 2016;27(11):3253–65. Epub 20160802. doi: 10.1681/asn.2016010098. PubMed PMID: 27486138; PubMed Central PMCID: PMCPMC5084898.

27. Sedgwick P. Multiple hypothesis testing and Bonferroni’s correction. Bmj. 2014;349:g6284. Epub 20141020. doi: 10.1136/bmj.g6284. PubMed PMID: 25331533.

28. Middeldorp JM, Brink AA, van den Brule AJ, Meijer CJ. Pathogenic roles for Epstein-Barr virus (EBV) gene products in EBV-associated proliferative disorders. Crit Rev Oncol Hematol. 2003;45(1):1–36. doi: 10.1016/s1040-8428(02)00078-1. PubMed PMID: 12482570.

29. Zhang Q, Holley-Guthrie E, Ge JQ, Dorsky D, Kenney S. The Epstein-Barr virus (EBV) DNA polymerase accessory protein, BMRF1, activates the essential downstream component of the EBV oriLyt. Virology. 1997;230(1):22–34. doi: 10.1006/viro.1997.8470. PubMed PMID: 9126259.

30. Dooley MM, de Gannes SL, Fu KA, Lindsey JW. The increased antibody response to Epstein-Barr virus in multiple sclerosis is restricted to selected virus proteins. J Neuroimmunol. 2016;299:147–51. Epub 20160831. doi: 10.1016/j.jneuroim.2016.08.016. PubMed PMID: 27725113.

31. Huang J, Tengvall K, Bomfim Lima I, Hedström AK, Butt J, Brenner N, et al. Genetics of immune response to Epstein-Barr virus: prospects for multiple sclerosis pathogenesis. Brain. 2024. Epub 20240417. doi: 10.1093/brain/awae110. PubMed PMID: 38630618.

32. Domínguez-Mozo MI, López-Lozano L, Pérez-Pérez S, García-Martínez Á, Torrejón MJ, Arroyo R, Álvarez-Lafuente R. Epstein-Barr Virus and multiple sclerosis in a Spanish cohort: A two-years longitudinal study. Front Immunol. 2022;13:991662. Epub 20220914. doi: 10.3389/fimmu.2022.991662. PubMed PMID: 36189297; PubMed Central PMCID: PMCPMC9515943.

33. Bray PF, Luka J, Bray PF, Culp KW, Schlight JP. Antibodies against Epstein-Barr nuclear antigen (EBNA) in multiple sclerosis CSF, and two pentapeptide sequence identities between EBNA and myelin basic protein. Neurology. 1992;42(9):1798–804. doi: 10.1212/wnl.42.9.1798. PubMed PMID: 1381067.

34. Lanz TV, Brewer RC, Ho PP, Moon JS, Jude KM, Fernandez D, et al. Clonally expanded B cells in multiple sclerosis bind EBV EBNA1 and GlialCAM. Nature. 2022;603(7900):321–7. Epub 20220124. doi: 10.1038/s41586-022-04432-7. PubMed PMID: 35073561; PubMed Central PMCID: PMCPMC9382663.

35. Kyllesbech C, Trier N, Slibinskas R, Ciplys E, Tsakiri A, Frederiksen JL, Houen G. Virus-specific antibody indices may supplement the total IgG index in diagnostics of multiple sclerosis. J Neuroimmunol. 2022;367:577868. Epub 20220418. doi: 10.1016/j.jneuroim.2022.577868. PubMed PMID: 35477126.

36. Lindsey JW. Antibodies to the Epstein-Barr virus proteins BFRF3 and BRRF2 cross-react with human proteins. J Neuroimmunol. 2017;310:131–4. Epub 20170720. doi: 10.1016/j.jneuroim.2017.07.013. PubMed PMID: 28778437.

37. Kuhlenbäumer G, Hannibal MC, Nelis E, Schirmacher A, Verpoorten N, Meuleman J, et al. Mutations in SEPT9 cause hereditary neuralgic amyotrophy. Nat Genet. 2005;37(10):1044–6. Epub 20050925. doi: 10.1038/ng1649. PubMed PMID: 16186812.

38. Mao H, Liu J, Shi W, Huang Q, Xu X, Ni L, et al. The expression patterns of Septin-9 after traumatic brain injury in rat brain. J Mol Neurosci. 2013;51(2):558–66. Epub 20130523. doi: 10.1007/s12031-013-0024-6. PubMed PMID: 23700216.

39. Tengvall K, Huang J, Hellström C, Kammer P, Biström M, Ayoglu B, et al. Molecular mimicry between Anoctamin 2 and Epstein-Barr virus nuclear antigen 1 associates with multiple sclerosis risk. Proc Natl Acad Sci U S A. 2019;116(34):16955–60. Epub 20190802. doi: 10.1073/pnas.1902623116. PubMed PMID: 31375628; PubMed Central PMCID: PMCPMC6708327.

40. Sherbet GV. Molecular Approach to Targeted Therapy for Multiple Sclerosis. CNS Neurol Disord Drug Targets. 2016;15(1):20–34. doi: 10.2174/1871527315666151110125840. PubMed PMID: 26560895.

41. Moreno MA, Or-Geva N, Aftab BT, Khanna R, Croze E, Steinman L, Han MH. Molecular signature of Epstein-Barr virus infection in MS brain lesions. Neurol Neuroimmunol Neuroinflamm. 2018;5(4):e466. Epub 20180607. doi: 10.1212/nxi.0000000000000466. PubMed PMID: 29892607; PubMed Central PMCID: PMCPMC5994704.

42. Osipova-Goldberg HI, Turchanowa LV, Adler B, Pfeilschifter JM. H2O2 inhibits BCR-dependent immediate early induction of EBV genes in Burkitt’s lymphoma cells. Free Radic Biol Med. 2009;47(8):1120–9. Epub 20090621. doi: 10.1016/j.freeradbiomed.2009.06.019. PubMed PMID: 19540913.

43. Maple PAC, Gran B, Tanasescu R, Pritchard DI, Constantinescu CS. An Absence of Epstein-Barr Virus Reactivation and Associations with Disease Activity in People with Multiple Sclerosis Undergoing Therapeutic Hookworm Vaccination. Vaccines (Basel). 2020;8(3). Epub 20200828. doi: 10.3390/vaccines8030487. PubMed PMID: 32872342; PubMed Central PMCID: PMCPMC7564729.

44. Torkildsen Ø, Nyland H, Myrmel H, Myhr KM. Epstein-Barr virus reactivation and multiple sclerosis. Eur J Neurol. 2008;15(1):106–8. Epub 20071127. doi: 10.1111/j.1468-1331.2007.02009.x. PubMed PMID: 18042233.

45. Yea C, Tellier R, Chong P, Westmacott G, Marrie RA, Bar-Or A, Banwell B. Epstein-Barr virus in oral shedding of children with multiple sclerosis. Neurology. 2013;81(16):1392–9. Epub 20130906. doi: 10.1212/WNL.0b013e3182a841e4. PubMed PMID: 24014504; PubMed Central PMCID: PMCPMC3806908.

46. Massa J, Munger KL, O’Reilly EJ, Falk KI, Ascherio A. Plasma titers of antibodies against Epstein-Barr virus BZLF1 and risk of multiple sclerosis. Neuroepidemiology. 2007;28(4):214–5. Epub 20070911. doi: 10.1159/000108113. PubMed PMID: 17851260.

47. Masuda S, Mori M, Arai K, Uzawa A, Muto M, Uchida T, et al. Epstein-Barr virus persistence and reactivation in neuromyelitis optica. J Neurol Neurosurg Psychiatry. 2015;86(10):1137–42. Epub 20141128. doi: 10.1136/jnnp-2014-308095. PubMed PMID: 25433035.

48. Shi Z, Kong L, Wang R, Wang X, Wang Z, Luo W, et al. Cytomegalovirus and Epstein-Barr virus infections in patients with neuromyelitis optica spectrum disorder. J Neurol. 2024. Epub 20240724. doi: 10.1007/s00415-024-12571-2. PubMed PMID: 39046523.

49. Graves J, Grandhe S, Weinfurtner K, Krupp L, Belman A, Chitnis T, et al. Protective environmental factors for neuromyelitis optica. Neurology. 2014;83(21):1923–9. Epub 20141022. doi: 10.1212/wnl.0000000000001001. PubMed PMID: 25339213; PubMed Central PMCID: PMCPMC4248458.

50. Merelli E, Bedin R, Sola P, Barozzi P, Mancardi GL, Ficarra G, Franchini G. Human herpes virus 6 and human herpes virus 8 DNA sequences in brains of multiple sclerosis patients, normal adults and children. J Neurol. 1997;244(7):450–4. doi: 10.1007/s004150050121. PubMed PMID: 9266465.

51. Nora-Krukle Z, Chapenko S, Logina I, Millers A, Platkajis A, Murovska M. Human herpesvirus 6 and 7 reactivation and disease activity in multiple sclerosis. Medicina (Kaunas). 2011;47(10):527–31. PubMed PMID: 22186115.

52. Khalesi Z, Tamrchi V, Razizadeh MH, Letafati A, Moradi P, Habibi A, et al. Association between human herpesviruses and multiple sclerosis: A systematic review and meta-analysis. Microb Pathog. 2023;177:106031. Epub 20230210. doi: 10.1016/j.micpath.2023.106031. PubMed PMID: 36775211.

53. Thakolwiboon S, Zhao-Fleming H, Karukote A, Pachariyanon P, Williams HG, Avila M. Regional differences in the association of cytomegalovirus seropositivity and multiple sclerosis: A systematic review and meta-analysis. Mult Scler Relat Disord. 2020;45:102393. Epub 20200712. doi: 10.1016/j.msard.2020.102393. PubMed PMID: 32683307.

54. Heerlein K, Jarius S, Jacobi C, Rohde S, Storch-Hagenlocher B, Wildemann B. Aquaporin-4 antibody positive longitudinally extensive transverse myelitis following varicella zoster infection. J Neurol Sci. 2009;276(1-2):184–6. Epub 20080920. doi: 10.1016/j.jns.2008.08.015. PubMed PMID: 18805556.

55. Sellner J, Hemmer B, Mühlau M. The clinical spectrum and immunobiology of parainfectious neuromyelitis optica (Devic) syndromes. J Autoimmun. 2010;34(4):371–9. Epub 20091022. doi: 10.1016/j.jaut.2009.09.013. PubMed PMID: 19853412.

56. Machado C, Amorim J, Rocha J, Pereira J, Lourenço E, Pinho J. Neuromyelitis optica spectrum disorder and varicella-zoster infection. J Neurol Sci. 2015;358(1-2):520–1. Epub 20151004. doi: 10.1016/j.jns.2015.09.374. PubMed PMID: 26440423.

57. Wang L, Zhou L, ZhangBao J, Huang W, Tan H, Fan Y, et al. Causal associations between prodromal infection and neuromyelitis optica spectrum disorder: A Mendelian randomization study. Eur J Neurol. 2023;30(12):3819–27. Epub 20230816. doi: 10.1111/ene.16014. PubMed PMID: 37540821.

